# On the sensitivity of non-pharmaceutical intervention models for SARS-CoV-2 spread estimation

**DOI:** 10.1101/2020.06.10.20127324

**Authors:** Kristian Soltesz, Fredrik Gustafsson, Toomas Timpka, Joakim Jaldén, Carl Jidling, Albin Heimerson, Thomas B. Schön, Armin Spreco, Joakim Ekberg, Örjan Dahlström, Fredrik Bagge Carlson, Anna Jöud, Bo Bernhardsson

## Abstract

**Introduction:** A series of modelling reports that quantify the effect of non-pharmaceutical interventions (NPIs) on the spread of the SARS-CoV-2 virus have been made available prior to external scientific peer-review. The aim of this study was to investigate the method used by the Imperial College COVID-19 Research Team (ICCRT) for estimation of NPI effects from the system theoretical viewpoint of model identifiability.

**Methods:** An input-sensitivity analysis was performed by running the original software code of the systems model that was devised to estimate the impact of NPIs on the reproduction number of the SARS-CoV-2 infection and presented online by ICCRT in *Report 13* on March 30 2020. An empirical investigation was complemented by an analysis of practical parameter identifiability, using an estimation theoretical framework.

**Results:** Despite being simplistic with few free parameters, the system model was found to suffer from severe input sensitivities. Our analysis indicated that the model lacks practical parameter identifiability from data. The analysis also showed that this limitation is fundamental, and not something readily resolved should the model be driven with data of higher reliability.

**Discussion:** Reports based on system models have been instrumental to policymaking during the SARS-CoV-2 pandemic. With much at stake during all phases of a pandemic, we conclude that it is crucial to thoroughly scrutinise any SARS-CoV-2 effect analysis or prediction model prior to considering its use as decision support in policymaking. The enclosed example illustrates what such a review might reveal.

## Introduction

The spread of the SARS-Cov-2 virus has challenged the world and political decisions are continuously made to reduce the virus’ s effect on population health and on health care organisations. In the absence of a vaccine, European populations can only safeguard themselves through behavioural measures ranging from increased personal hygiene to social distancing. Currently, policymakers are therefore seeking scientific guidance upon which to base national exit strategies from non-pharmaceutical interventions (NPIs) in a safe, yet efficient manner.

A series of reports have presented estimated effects of various NPIs on the spread of SARS-CoV-2 infection, such as case isolation, closure of schools, restrictions specifically towards vulnerable groups such as people aged over 70 years, or a total lockdown. Specifically, a series of reports set out to quantify the effect of NPIs [1,2]. These reports have since been widely used as support for political decisions that aim at reducing the rate at which the virus spreads [3].

### The Imperial College reports

A scenario analysis, *Report 9* [1], released online March 16, 2020, by the *Imperial College COVID-19 Response Team* (ICCRT) was influential in the change of the COVID-19 response policy in the UK [3] and is likely to have affected policy decisions in other European countries.

In a subsequent document, *Report 13* [2], released online March 30, a method was presented to estimate the actual effects of different NPIs that had been effectuated by then in 11 European countries. Based on effectuation dates of the NPIs and available time series of mortalities in each country, the influence of individual NPIs on the reproduction number *R*_*t*_ of SARS-CoV-2 infection was estimated from data.

The system model in *Report 13*, relating the effect of NPIs to *R*_*t*_, could if reliable be very useful when weighing their efficiency versus cost for society. The method upon which it is based has, however, not yet been thoroughly and externally peer-reviewed.

### Objectives

In this study we investigate the method used in ICCRT *Report 13* for estimation of NPI effects from the system theoretical viewpoint of model identifiability. In particular, we examine input sensitivities and how identifiability issues can influence the results.

Our primary purpose is not to offer a critique of Report 13. The ambitions of that report, and the release of the supporting data and models into the public domain, are to be commended. Rather our objective is to illustrate how structural assumptions that are inevitably built into pandemic models add to the sensitivity of their estimations and predictions.

### Pandemic models

Typical for a pandemic is that the quality of data on infected individuals and deaths is initially limited by inconsistent detection of cases, reporting delays, and poor documentation. It is therefore understandable that data has been the main focus in the debate surrounding pandemic estimation and prediction models, but this has overshadowed the discussion about the models themselves.

*Report 9* used a *mechanistic* model to represent disease spread at a local level, considering social interactions that could produce transmission events. It models the effects of 5 NPIs: *case isolation, closure of schools and universities, social distancing for those over age 70, voluntary home quarantine*, and *social distancing of the entire population* (lockdown).

The *Report 9* model was parameterised using results from previous studies and used “plausible and largely conservative (i.e. pessimistic) assumptions about the impact of each intervention”. A representative such assumption is that the closing of schools eliminates infections in the schools but increases contact rates within affected families by a certain factor, and in the community in general by some other factor. When supported by data, *mechanistic models* such as the one of *Report 9* can be used to explain and understand the evolution of a pandemic and how it is affected by modelled interventions. Counterfactual (“what-if?”) analysis can be done safely within the model, and the results of possible interventions can be estimated.

However, using a mechanistic approach during the initial stage of a pandemic, with unknown social parameters and mechanisms of infection, is a challenging task. The sizable number of parameters that are difficult to determine are likely to make predictions uncertain [4,5].

In contrast to the mechanistic system model in *Report 9*, the system model in *Report 13* is to a larger extent *phenomenological* and uses a data-driven approach, rather than detailed underlying mechanisms, to determine input-output relationships. Its parameterisation is limited to country-specific estimates of the infection fatality ratio, a serial interval distribution assumed to be equal in all countries, and multiplicative factors describing how individual NPIs affect *R*_*t*_. Instead of assuming that the effect of individual NPIs is known, as was done in *Report 9*, the modelling aim is to identify these factors from mortality data.

Similar analyses, estimating the effect of NPIs effectuated in Wuhan, have also been presented in [6-7], albeit using other methods and additional information such as mobility data, detected cases, and reports of symptom onset. On May 4, ICCRT also released *Report 20* [8], in which the *Report 13* system model had been extended to include mobility data. It is described that this extended model can be used to “calculate the deaths averted by keeping mobility at current levels [in Italy]”.

### Estimating the impact of NPIs

In *Report 13*, five NPIs are defined, see Figure 1a. When effectuated, each NPI is assumed to instantaneously result in a step-change in *R*_*t*_, as illustrated in Figure 1b, reproducing results from *Report 13*.

**Figure 1.**
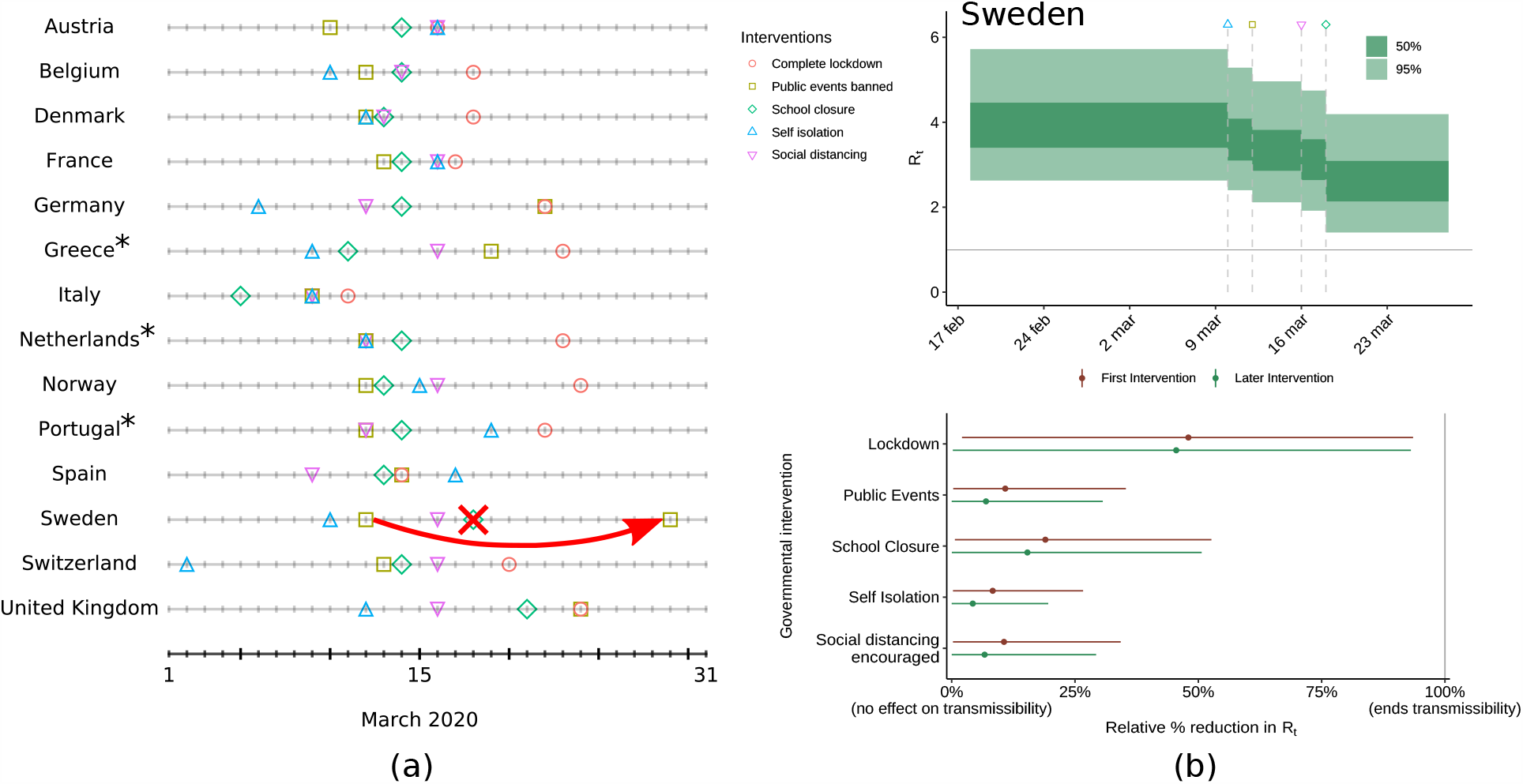
**(a)** Dates that NPIs were effectuated in individual countries, according to [9]. The annotations illustrate code revisions introduced by the Imperial College modellers following their release of *Report 13* [2]: On April 9, the date on which *public events* were banned in Sweden was revised by the modellers in [9]. In the same code revision, *school closure* was no longer defined to have taken place in Sweden. **(b)** Diagrams showing reproduced results from [2]. *On April 6, the model code [9] was extended to include three additional countries.

Figure 2 provides a high-level view of the model used in *Report 13*.

**Figure 2.**
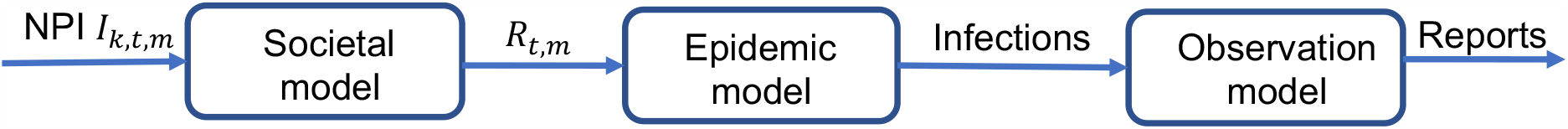
The left block models how NPI *k* at time *t* affects the reproduction number *R*_*t,m*_ in country *m*. The middle block includes a model for how new infections evolve as a function of the effective reproduction number, while the right block models how reported deaths depend on previous infections.

The *societal model* describes how different NPIs affect the reproduction number. It assumes that *R*_*t*_ is only affected by the NPI parameters, and by herd-immunity effects resulting from a diminishing susceptible-to-infected ratio. Data from different countries are effectively pooled through the assumption that the NPI parameters are not country-specific: the factor of relative change in *R*_*t*_ resulting from effectuating a particular NPI is the same, regardless of in which country the NPI is effectuated. The factors are assumed time-independent, with the exception that a bonus factor is ascribed to an NPI if it is the first to be effectuated in a country. This is illustrated in the bottom pane of Figure 1b.

Some country-specific flexibility is, however, provided through the basic reproduction number *R*_*0*_ being modelled as a country-specific parameter, co-identified with the pooled parameters affecting *R*_*t*_. In a later version of the model [9], released April 24 (after *Report 13*), the *lockdown* NPI, being the last NPI to be effectuated in all countries, except for Sweden, was also ascribed some (practically rather limited) country-specific flexibility.

The *epidemic model*, which can be interpreted as an SIR type model [10], describes how new infections evolve as a function of the reproduction number.

The *observation model* tries to explain how reported deaths depend on previous infections and is implemented by a stochastic infection-to-death distribution with mean value 23.9 days and an infection fatality ratio, both based on previous reports [11].

The combined model is compared with daily reported deaths (source: ECDC [12]), and a Bayesian framework is employed to compute estimates of the pooled NPI parameters, along with the country-specific *R*_*0*_ values. In *Report 13* estimation results are reported using 50 % and 95 % credible intervals.

## Methods

A re-analysis of the system model used by the ICCRT for the estimation of NPI effects was performed using the published code [9]. The data used for our analysis was collected from the original source (ECDC data repository [12]). Our re-analysis consists of three parts: First, we perform an NPI sensitivity analysis. Second, we study the importance of lockdown. Third, we perform a model identifiability analysis.

### NPI sensitivity analysis

A challenge in the modelling of NPI effects is to accurately represent when and to what extent a particular NPI was effectuated. We investigate two case examples:

- Higher education instances in Sweden transitioned to online teaching on March 18, while elementary schools remained open. The version of the model used in *Report 13* [2] (dated March 30), defined *school closure* to have taken place on March 18. However, in the revised model [9] released April 24, *school closure* in Sweden had been re-defined not to have taken place.
- Similarly, public events exceeding 500 persons were banned in Sweden on March 12. On March 29, the restriction was tightened to 50 persons. *Report 13* defined the date of the *public events* ban to March 12; in [9] this date had been revised to March 29.

### Estimating the importance of lockdown

The results from our initial sensitivity analysis (Figure 3, further below) indicated that the estimated importance of *lockdown* had increased with an additional month of data available since the release of *Report 13*. It was also observed that the model displayed difficulties in fitting its results to the data for Sweden. With 14 countries in the analysis, Sweden was still the only country without the NPI *lockdown* being effectuated. To give the model a chance to evaluate the efficiency of the *lockdown* intervention on a more equal footing, we ran the code on input data from only two countries: The UK and Sweden.

**Figure 3.**
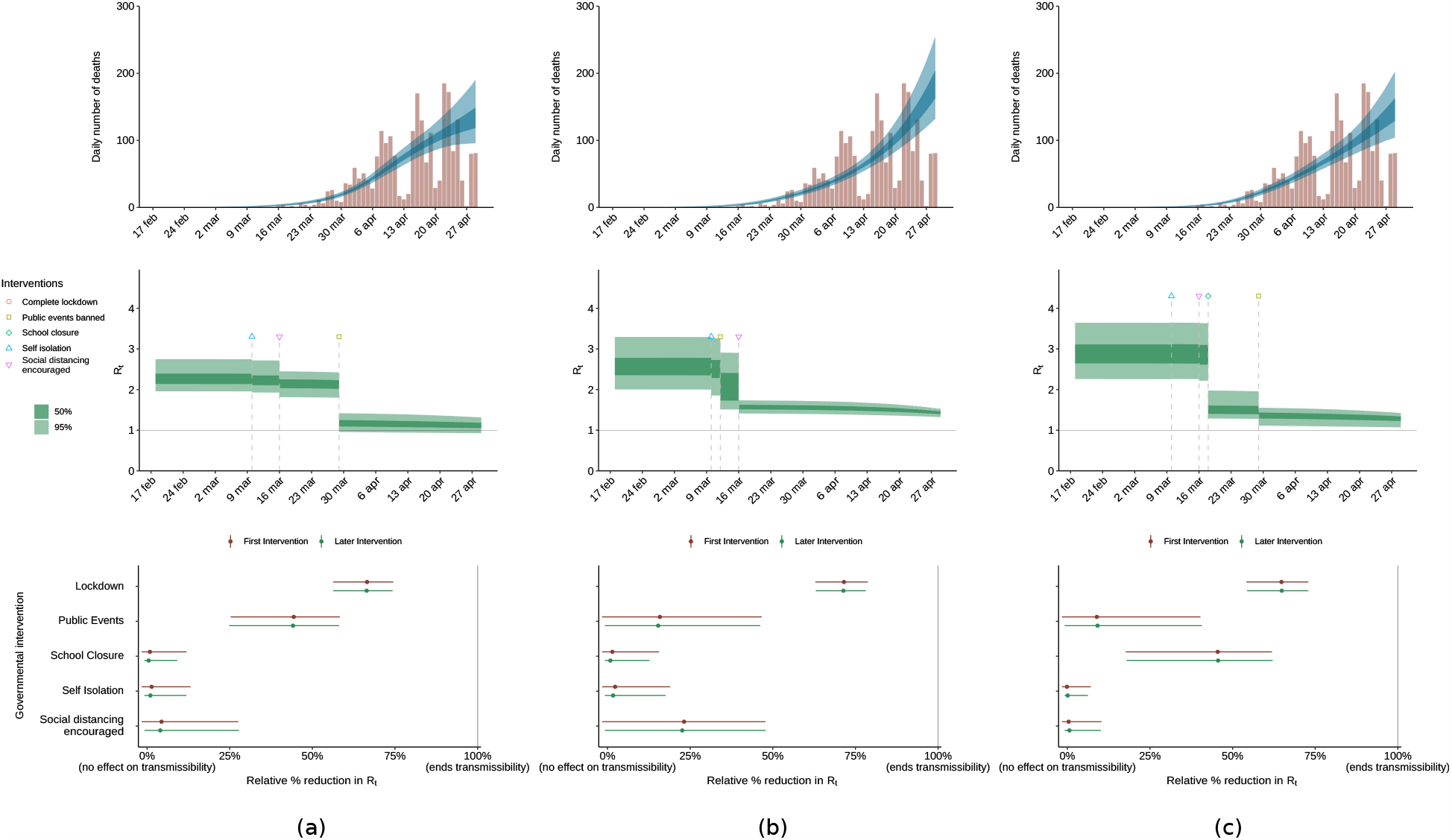
Each column corresponds to one execution of the code [9]: **(a)** Definitions according to the April 29 version of the Imperial College model [9], (*Public events* ban in Sweden March 29; *school closure* defined to not have taken place). **(b)** Same as (a), but with the *public events* ban moved back to March 12 as in [2]. **(c)** Same as (a), but with *school closure* defined to have taken place March 18 as in [2]. From top down: daily reported deaths (source: ECDC) and model fit; estimate of the reproduction number *R*_*t*_. The bottom row shows the impact of each intervention category on *R*_*t*_, affecting all the 14 considered countries. The large effects of redefining a single intervention date in one country indicate that the NPI impact cannot be reliably estimated from the available data. Code reproducing the results is available through [14]. See [2] for details on how to interpret the diagrams.

### Model identifiability analysis

To complement the empirical results and study the NPI parameter sensitivity, we then considered an idealised hypothetical case where we assumed that one over the relevant period could measure the country-specific reproduction number *R*_*t,m*_ directly in each country. In estimation-theoretic terms, the block diagram in Figure 2 forms a Markov chain. This implies that direct access to *R*_*t,m*_ obviates the need to access the original data, and a study of parameters identifiability given *R*_*t,m*_ provides provably optimistic conclusions regarding the NPI parameter identifiability. Conversely, if NPI parameters are hard to identify given *R*_*t,m*_, they are even harder to identify from mortality data.

The estimation of the NPI parameters and the country-specific basic reproduction number from *R*_*t,m*_ via the societal model can be expressed as a standard linear regression problem after taking logarithms, due to the multiplicative nature of the NPI effects:

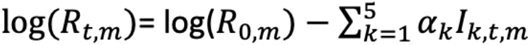

Here *I*_*k,t,m*_ is an indicator variable with value 0 or 1, that signals if NPI *k* was in effect in country *m* on day *t* and *α*_*k*_ is a parameter that models the effectiveness for NPI *k*, using the notation of [2]. This allows us to determine the difficulty of estimating the NPI parameters *α*_*k*_ and log(*R*_*0,m*_) by studying the *design* or *regression* matrix of the linear regression problem [13]. We complement this study by illustrating how the country-specific *R*_*0,m*_ are used to account for model mismatch when more data becomes available, by comparing the estimated *R*_*0,m*_ parameter in the full *Report 13* model when applied to data up until March 28 and up until April 29.

### Model code versions

To investigate the effects of NPI definitions we executed the code of the model with the two minor modifications given by reversing the changes of dates described above. The basis for the production of our results was the model code made available on Github by the ICCRT [9]. We created a fork [14] of the code, based on the April 29 commit da9a8a9 of [9]. Below we reference the specific commits of this fork [14], used to obtain our results. The model was executed in the “full” mode, according to the README instructions provided with the April 29 version of [9].

The reproduction of results from *Report 13* shown in Figure 1b was obtained using commit 58df932 of [14], being equivalent to the March 29 version of [9], except for a plotting script copied from the April 29 version. Mortality data, provided with the code and originating from ECDC [12], was used.

The remainder of our results were obtained using code based on the April 29 version of [9]. In order to investigate the effect of NPI definitions, the changes annotated in Figure 1a were implemented in commit 49bd665 of [14]. Mortality data available on April 29 was retrieved from ECDC [12]. This setup was used to generate Figure 3.

In order to investigate how the pooling of countries affects the estimated relative importance of NPIs, the April 29 version of [9] was modified by removal of all countries except Sweden and the UK. This was implemented in commit 38c9057 of [14]. Mortality data available on April 29 was retrieved from ECDC [12] also for this investigation. This setup was used to generate Figures 4 and 5.

**Figure 4.**
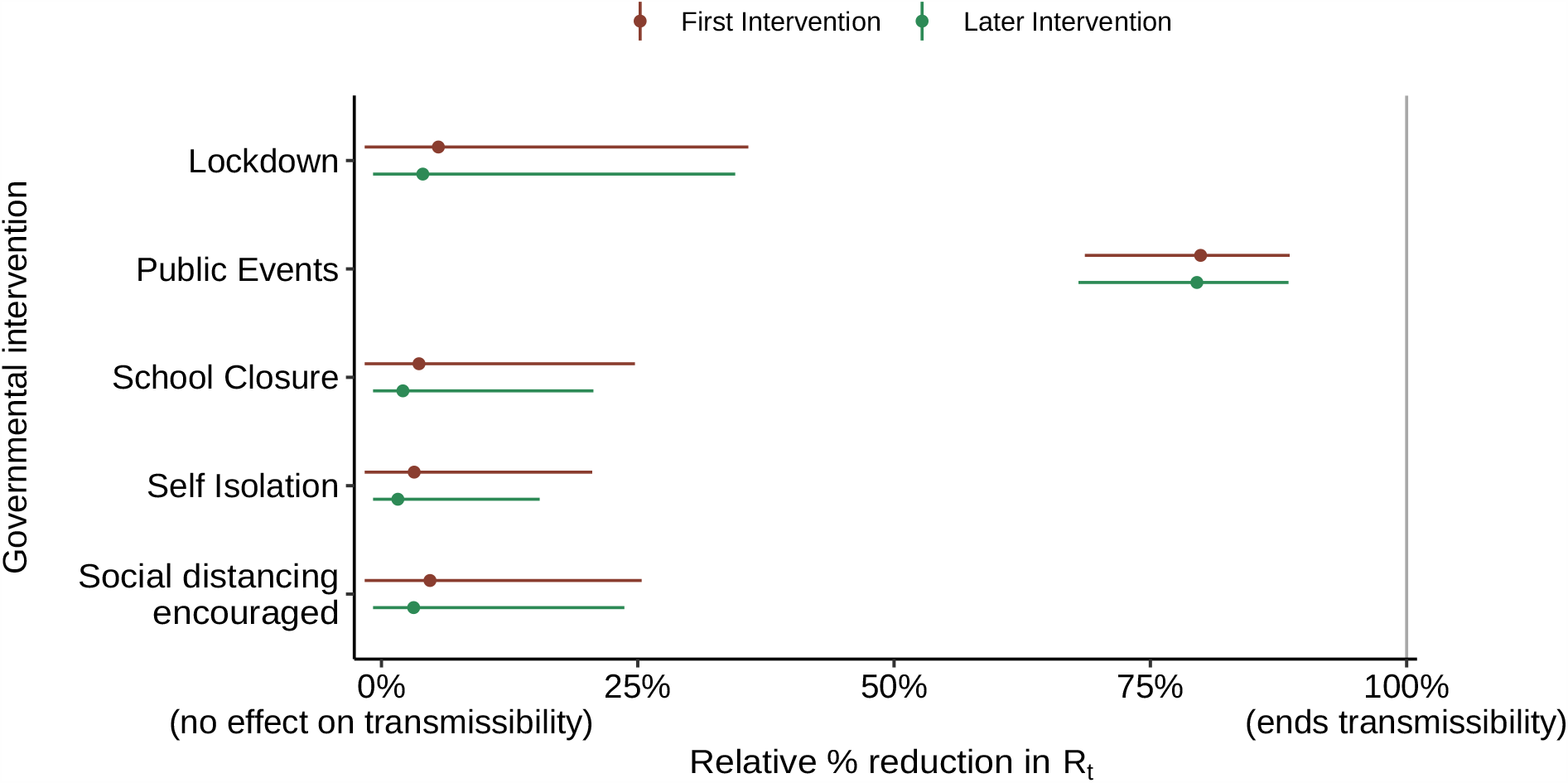
Results using [9], configured to estimate the efficiency of NPIs based on reported mortality data (source: ECDC [12]) from the UK and Sweden up to and including April 29. The model now estimates that *public events* ban has been the most efficient intervention, while the importance of *lockdown* being small

**Figure 5.**
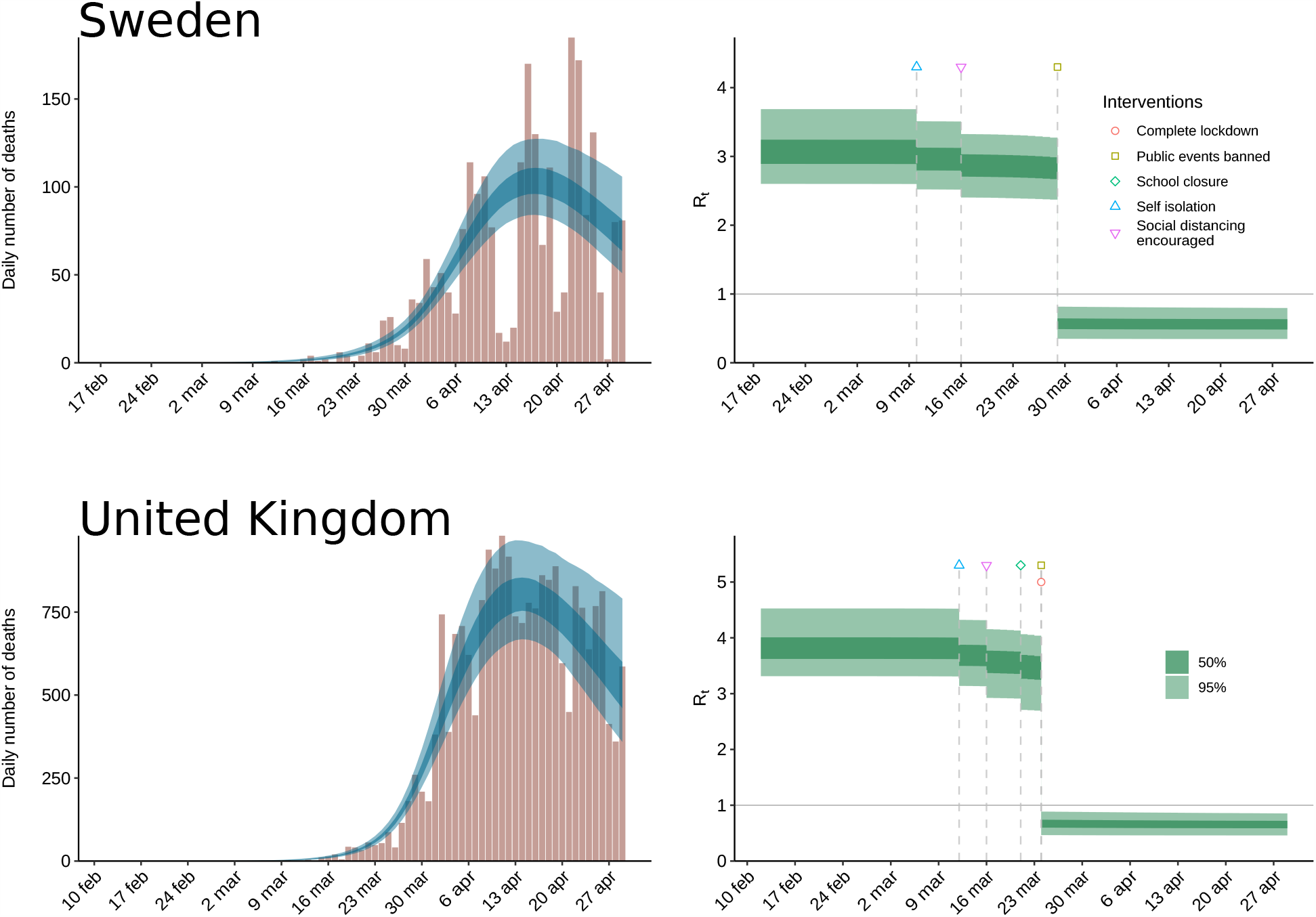
Results using [9], configured to estimate the efficiency of NPIs based on reported mortality data (source: ECDC [12]) from the UK and Sweden up to and including April 29. The reduction of *R*_*t*_ is almost exclusively ascribed to the *public events* ban NPI; the model estimates the importance of *lockdown* to be negligible. The model estimates *R*_*t*_ to have dropped below 1 at the end of March in both countries, with stated credibility exceeding 95 %.

In order to compare *R*_*0*_ estimates provided by the model, when driven by data available by different dates, the April 29 version of [9] was run with ECDC data available on March 28, and the corresponding data available on April 29. This was implemented in commit 895e1fd of [14] and used to generate Figure 6.

**Figure 6.**
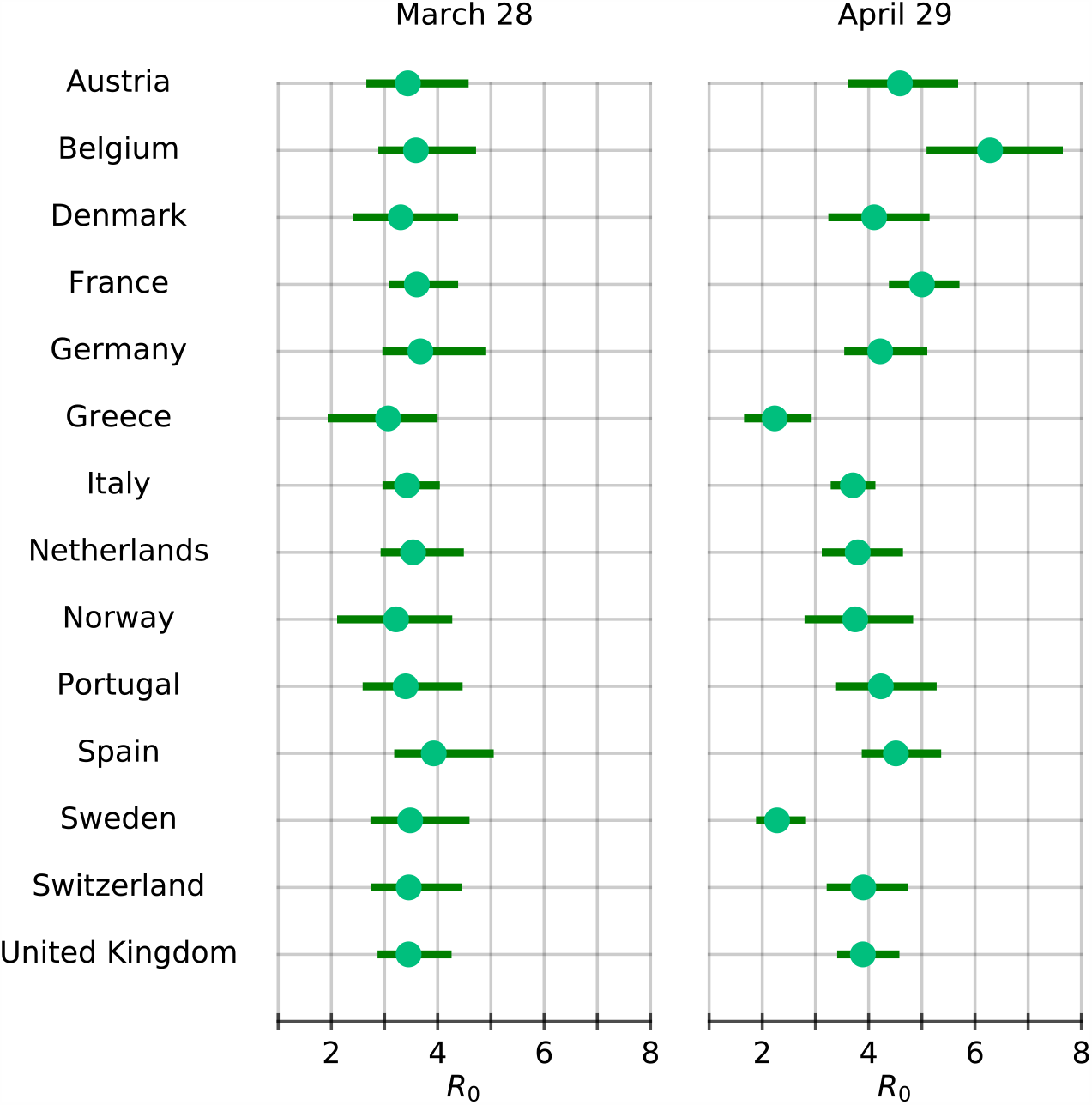
Country-specific estimates of *R*_*0*_ produced using [9]. To the left: using mortality data up to and including March 28 as in [2]. To the right: using data up to and including April 29 (data source: ECDC [12]).

## Results

### NPI sensitivities

Our analysis of *Report 13* [2] showed that also a minor change in a single NPI definition in one country can result in large changes of the estimated NPI effects on *R*_*t*_ in all 14 European countries. Figure 1a shows the revisions between [2] and the April 29 release of [9] for the two NPIs mentioned above. Figure 3 illustrates the result of executing three instances of the model, to investigate the consequences of the seemingly subtle changes in NPI definitions introduced by the Imperial College modellers. The main results, the estimated NPI parameters, are given in the bottom pane.

Compared to the original settings, moving the *public events* ban date in Sweden to March 12 resulted in the model estimating a *public events* ban to have a decisively lower impact on *R*_*t*_, not only in Sweden but in *all* 14 European countries. Correspondingly, redefining *school closure* to have taken place in Sweden resulted in the model estimating *school closure* to have a much larger impact on *R*_*t*_, again in all countries. There are also noticeable changes in the claimed certainty of the estimated NPI parameters.

### Estimated the importance of lockdown

Pooling Sweden and UK data resulted in major reattributions of NPI effects, as shown in Figures 4 and 5. The system model now attributes the reduction in *R*_*t*_ to the *public events* ban and estimates a negligible effect of the *lockdown* NPI.

### Model identifiability

The regression matrix for the NPI parameters *α*_*k*_ and log(*R*_*0,m*_) was found to be rather ill-conditioned, with a condition number of 20 when computed over *R*_*t,m*_ corresponding to the whole month of March during which the interventions took place. Further analysis revealed that certain combinations of the parameters are more accurately estimated than their individual values.

Thus, there is a high correlation between some of the parameters which cannot be seen in the confidence intervals shown in Figures 1, 3, 4 and 6, or any of the plots shown in [2] involving individual parameter standard deviations. The relatively poor conditioning results from all interventions taking place during a short time period. This analysis can be performed without having any mortality data at hand. The results only depend on the model assumption and the input data, i.e. the dates the NPIs were effectuated.

As the regression matrix is of full rank, it follows that the NPI parameters are structurally identifiable from *R*_*t,m*_, and are likely structurally identifiable [15] from the mortality data in theory.

However, the difficulty of practically identifying model parameters of even the simplest epidemic models from data [15] indicates that it is difficult to obtain reliable estimates of *R*_*t,m*_ during a pandemic, and any errors made in estimating *R*_*t,m*_ are amplified in the estimate of the NPI effect estimates *α*_*k*_ due to the ill-conditioned regression matrix. The problem is exacerbated in practice by varying standards of reporting mortality data.

Since all countries, except Sweden, were defined to have effectuated all 5 NPIs, the methodology in [2] is forced to give similar relative reduction from the initial *R*_*0*_ value to the final value of *R*_*t*_. Figure 6 shows the *R*_*0*_ estimates for the 14 countries, produced using [9] driven by data available March 28 (left), and April 29 (right), respectively.

The March 28 data, corresponding to the early stage of the pandemic, results in only small inter-country variations in the *R*_*0*_ estimates, compared to those obtained using the April 29 data. An explanation of the large variations seen in the April 29 case can be that forcing combined NPI effects to be the same in all countries, except Sweden, offers *R*_*0*_ as the only remaining free parameter to effectively explain any country-wise variations in the final *R*_*t*_,. The relative impact of the NPIs on *R*_*t*_ is the same in all countries, and when this does not fit the data, the model has to compensate by adjusting the country-specific *R*_*0*_ estimates.

## Discussion

The aim of this study was to investigate the method used in ICCRT *Report 13* for estimation of NPI effects from the system theoretical viewpoint. We found the combined effects of high input sensitivity, and the assumption of *R(t)* being driven solely by the NPIs, to constitute a fundamental limitation which should be considered when the modelling results are used as a basis for policymaking.

It is hard to judge whether, for example, a partial transition to online teaching constitutes a *school closure* or not. Similarly, the crowd size limit associated with the *public events* ban NPI remains a free design parameter for the modeller to decide. The point is not to argue if a school closure took place or not, or what the most appropriate crowd size limit is. Instead, our findings highlight remarkably large effects resulting from minor changes in input data. If the NPI modelling results are to be used as support for policy decisions, it is not acceptable that subtle interpretations of NPI definitions in a single small country, like those reported here, have a pivotal impact on the estimated intervention effects in all 14 modelled countries. It is hence inherently difficult to confidently ascribe changes in *R*_*t*_ to specific NPIs that jointly took place over a matter of days, based on data with inconsistent detection and reporting of cases.

Since the effect of each NPI is modelled by a multiplicative factor applied to *R*_*t*_, an NPI that is deemed 75 % effective reduces *R*_*t*_ by a multiplicative factor of 1-0.75=0.25. A country that effectuates two NPIs with 50 % effectiveness on the same day will, therefore, see exactly the same reduction in *R*_*t*_ as a country with a single 75 % effective NPI, as 0.5*0.5 = 0.25. The model can for this reason not distinguish between the two cases based on available data alone, no matter how good the data is. This effect is most apparent in the UK (Figure 5), which effectuated *lockdown* and *public events* ban on the same day. Based on UK data alone, the relative effectiveness of the *lockdown* and the *public events* ban are thus not even structurally identifiable, see [15] for a definition of this concept. Due to the pooling of NPI effectiveness across countries, and since Sweden did not effectuate *lockdown*, the model naturally explains the reduction in *R*_*t*_ observed in the data for both Sweden and the UK using the *public events* ban: The *lockdown* and *public events* ban are equally likely explanations for the reduction in *R*_*t*_ for the UK and the only explanation allowed by the model for the observed slowdown of deaths in Sweden.

The nominal case considered in *Report 13*, does not suffer from the pathological case of any NPI being effectuated on the same day across all 14 countries, see Figure 3, but the observed sensitivity issues can still be qualitatively understood from the analysis of the regression matrix for the linear regression problem at the heart of estimating the NPI parameters. The identifiability issues are acknowledged in the *Report 13* [2], followed by the statement that “while individual impacts cannot be determined, their estimated joint impact is strongly empirically justified”. It is easy to overlook this remark, as it might be overshadowed by the subsequent presentation of credible intervals for the effect of individual NPIs. As seen in Figure 3 these credible intervals shrink as more data becomes available, hiding the fundamental identifiability problems of the underlying model, giving a false sense of the reliability of the results. The issue is sharpened by the use of [9] and other non-validated models having been used as decision support for policymaking [3,5].

There are several other aspects of the model in [2] that deserve scrutiny. For instance, we would argue that it is fundamentally questionable to determine the joint impact of NPIs on *R*_*t*_ using a model based on the assumptions that changes in *R*_*t*_ are *de facto* only driven by the NPIs. Section 8.4.4 of [2] describes an attempt to assess this assumption using a Gaussian process as prior, but no results are presented^1^.

Using established principles from systems theory, we have demonstrated that even a seemingly simplistic and data-driven phenomenological model can suffer from severe input-sensitivity and identifiability issues. With much at stake during all phases of a pandemic, we conclude that it is crucial to thoroughly scrutinise any SARS-CoV-2 estimation or prediction model, prior to considering its use as decision support in policymaking. Such scrutiny relies on modellers following the practice used by the ICCRT in sharing open source code.

## Data Availability

All source code and data for full and completely transparent reproduction of all our results is available through https://github.com/albheim/covid19model

## Conflict of interest

None.

## Funding statement

This work was partially supported by the ELLIIT Strategic Research Area, by the Wallenberg AI, Autonomous Systems and Software Program (WASP) funded by the Knut and Alice Wallenberg Foundation, and by the Swedish Foundation for Strategic Research (SSF) via the project ASSEMBLE (contract number: RIT15-0012).

## Acknowledgements

We would like to acknowledge Ericsson Research, Lund Sweden, for hosting our model runs in their data centre.

## Footnotes

1 We have failed to get a response from the ICCRT when asking about material supporting the conclusions of Section 8.4.4.

## Notes

### Competing Interest Statement

The authors have declared no competing interest.

## References

[1] Ferguson NM, Laydon D, Nedjati-Gilani G, Imai N, Ainslie K, Baguelin M, et al. Report 9 - Impact of non-pharmaceutical interventions (NPIs) to reduce COVID-19 mortality and healthcare demand. [Preprint]. [posted 2020 March 16; cited 2020 April 29]. Available from: https://doi.org/10.25561/77482

[2] Flaxman S, Mishra S, Gandy A, Unwin HJT, Coupland H, Mellan TA, et al. Report 13 - Estimating the number of infections and the impact of non-pharmaceutical interventions on COVID-19 in 11 European countries. [Preprint]. [posted 2020 March 30; cited 2020 April 29]. Available from: https://doi.org/10.25561/77731

[3] Boseley S. New data, new policy: why UK’ s coronavirus strategy changed. The Guardian. 2020 March 16 [cited 2020 May 3]. Available from: https://www.theguardian.com/world/2020/mar/16/new-data-new-policy-why-uks-coronavirus-strategy-has-changed

[4] Timpka T, Eriksson H, Gursky EA, Nyce JM, Morin M, Jenvald J, et al. Population-based simulations of influenza pandemics: validity and significance for public health policy. Bull World Health Organ. 2009 Apr;87(4):305–11.

[5] Avery C, Bossert W, Clark A, Ellison G, Ellison S. Policy implications of models of the spread of coronavirus: perspectives and opportunities for economists. NBER Working Paper No. 27007 [Preprint]. 2020 [cited 2020 May 5]. Available from: https://doi.org/10.3386/w27007

[6] Lai S, Ruktanonchai NW, Zhou L, Prosper O, Luo W, Floyd JR, et al. Effect of non-pharmaceutical interventions to contain COVID-19 in China. Nature [Internet]. 2020 May [cited 2020 May 11]. Available from: https://www.nature.com/articles/s41586-020-2293-x.

[7] Leung K, Wu JT, Liu D, Leung GM. First-wave COVID-19 transmissibility and severity in China outside Hubei after control measures, and second-wave scenario planning: a modelling impact assessment. The Lancet. 2020 April 25;395(10233):1382–1393.

[8] Vollmer M, Mishra S, Unwin H, Gandy A, Melan T, Bradley V, et al. Report 20: A sub-national analysis of the rate of transmission of Covid-19 in Italy. [Preprint]. [posted 2020 May 4; cited 2020 May 10]. Available from: https://doi.org/10.25561/78677

[9] Imperial College London. covid19model [software]. [cited 2020 April 29]. Available from: https://github.com/ImperialCollegeLondon/covid19model

[10] Kermack WO, McKendrick AG. A contribution to the mathematical theory of epidemics. Proc. R. Soc. Lond. A. 1927 August 1;115(772):700–721.

[11] Verity R, Okell LC, Dorigatti I, Winskill P, Whittaker C, Natsuko I, et al. Estimates of the severity of COVID-19 disease. MedRxiv [Preprint]. [posted 2020 March 13; cited 2020 May 11]. Available from: http://medrxiv.org/lookup/doi/10.1101/2020.03.09.20033357 doi: https://doi.org/10.1101/2020.03.09.20033357

[12] European Center for Disease Prevention and Control (ECDC). Geographic distribution of COVID-19 cases worldwide. 2020 [cited 2020 Apr 29]. Available from: https://www.ecdc.europa.eu/en/publications-data/download-todays-data-geographic-distribution-covid-19-cases-worldwide.

[13] Ljung L. System identification: theory for the user. Prentice Hall PTR; 1999.

[14] Heimerson A. covid19model fork [software]. 2020. Available from: https://github.com/albheim/covid19model

[15] Tuncer N, Le TT. Structural and practical identifiability analysis of outbreak models. Mathematical biosciences. 2018;299:1–18.

